# SIEVE: Locus-Anchored Drug Prioritization for Complex Disorders

**DOI:** 10.64898/2026.04.15.26350958

**Authors:** Eric V. Strobl

## Abstract

**Motivation:** Complex disorders arise from multiple genetic mechanisms, but most drug-prioritization methods treat each disorder as a single phenotype and therefore miss locus-specific therapeutic opportunities.

**Results:** We present SIEVE, a framework that decomposes complex disorders into genetically localized subphenotypes and links GWAS summary statistics, reference expression, and perturbational transcriptional profiles to prioritize compounds that target locus-anchored disease mechanisms. SIEVE also constructs genetically calibrated mechanism vectors, projects away nonspecific expression programs using negative anchors, and aggregates evidence across cell lines, doses, and time points to produce robust drug rankings. Across simulations and analyses of real data, SIEVE improves compound prioritization relative to existing methods and shows that subphenotype-aware, genetics-guided modeling can sharpen therapeutic discovery in heterogeneous disorders.

**Availability and Implementation:** R implementation: github.com/ericstrobl/SIEVE.

## Introduction

Drug discovery for complex disorders remains challenging because clinical phenotypes often reflect multiple underlying biological processes and thus may not map cleanly onto a single therapeutic target (Sun et al., 2023). Consequently, many effective therapies act pleiotropically across several pathways, complicating the design of new compounds that precisely modulate the relevant pathways in the appropriate combination (Stefan and Rafehi, 2024; Munson et al., 2024). By contrast, Mendelian diseases are often more tractable because they allow more direct identification of causal targets (Lalagkas and Melamed, 2024). We therefore hypothesized that decomposing complex disorders into genetically more “Mendelian-like” components could uncover cleaner therapeutic targets and, in turn, facilitate more effective drug discovery.

Yet component decomposition alone does not guarantee accurate drug prioritization, because the molecular data used to link disease mechanisms to compounds are often noisy and confounded. In particular, large-scale perturbational gene expression datasets are affected by substantial technical variation, biological heterogeneity, and latent confounding (Musa et al., 2018). Moreover, each compound is profiled across multiple experimental contexts, including cell line, dose, and treatment duration, and the context combinations often differ across drugs (Wang and Novick, 2023). This structure makes drug prioritization challenging because signatures can vary substantially across contexts. In practice, noise often overwhelms signal, producing unstable drug scores and poorly calibrated rankings (Lim and Pavlidis, 2021).

The above challenges have motivated a broad class of compound-prioritization methods that focus primarily on denoising molecular measurements and ranking compounds according to how strongly a drug-induced perturbational profile reverses a disease-expression signature. For example, the original Connectivity Map (Lamb et al., 2006) and sscMap (Zhang and Gant, 2009) algorithms match ranked disease and drug signatures using prespecified up- and downregulated gene sets; NFFinder extends transcriptomic profile matching across multiple resources (Setoain et al., 2015); Cogena performs drug enrichment on co-expression modules rather than on flat signatures (Jia et al., 2016); DrInsight replaces fixed query signatures with gene-wise concordance statistics to identify concordantly expressed genes (Chan et al., 2019); metaLINCS aggregates replicate perturbation experiments to produce compound-level scores (Kwee et al., 2022); and Pathopticon incorporates cell-type-specific gene–perturbagen networks together with pathophenotypic congruity scores (Halu et al., 2025). Despite these methodological differences, these approaches remain fundamentally correlation-based: they prioritize compounds by how strongly drug and disease signatures oppose one another, without explicitly distinguishing downstream consequences of disease from causal drivers.

Recognizing this limitation, a second class of methods seeks to strengthen causal interpretation by anchoring disease signatures in human genetics. Rather than relying solely on observed disease expression, these approaches replace it with genetically predicted expression. For example, So et al. (2017) compared GWAS-imputed transcriptomes with drug-induced signatures from CMap in psychiatric disorders; Wu et al. (2022) used S-PrediXcan-derived disease signatures with iLINCS and then evaluated top candidates in electronic health record data; and TReD represents TWAS-derived disease signatures as multigene vectors, embeds them in the same space as LINCS drug-response profiles, and computes a reversal distance for each drug (He et al., 2025). These methods provide stronger mechanistic grounding because the disease signature is tied more directly to genetic risk. However, all of these methods still treat disease as a single aggregate phenotype. Thus, they improve the causal anchoring of compound prioritization without decomposing heterogeneous disorders into separable components.

Taken together, prior work addresses either noisy molecular data or causal ambiguity, but not both in a way that also confronts fundamental disease heterogeneity. In this paper, we address all three challenges by introducing **SIEVE** (**S**ubphenotype-**I**nformed **E**valuation of therapeutic **V**ectors from **E**xpression), a compound-prioritization framework outlined in Figure 1. SIEVE strengthens Mendelian-like signal while suppressing nuisance variation at multiple stages of the pipeline through complementary steps:

1. **Disease-level signal amplification**. SIEVE first reduces phenotypic heterogeneity by decomposing a complex disorder into a small set of Mendelian-like components, each of which aims to concentrate genetic association at a single causal locus. It then prioritizes therapeutics at the level of these Mendelianized, locus-anchored components rather than at the level of the original broad diagnosis.
2. **Genetically anchored deconfounding of reference expression**. SIEVE then uses the lead variants as an instrument set to extract the component of reference gene expression that is coupled to genetic variation. It subsequently calibrates this expression-derived mechanism toward the target using a strategy similar to two-stage least squares, thereby reducing the influence of latent confounding.
3. **Prior-guided noise reduction in reference and perturbational expression**. SIEVE further refines both the inferred mechanism vector and the perturbational signatures by incorporating prior information about drugs that are known to be ineffective or harmful, such as poisons and cytotoxic compounds. Because cellular toxicity can induce generic stress-response programs that make unrelated compounds appear spuriously similar at the transcriptional level (Szalai et al., 2019), SIEVE first identifies the expression directions associated with these negative controls and then projects both representations away from them. This projection suppresses irrelevant variation and sharpens mechanistically relevant signal.
4. **Robust drug scoring and aggregation**. SIEVE subsequently scores each perturbation by the cosine similarity between its expression signature and the inferred mechanism vector, thereby emphasizing directional concordance rather than absolute magnitude. It then aggregates condition-level scores to the drug level using gene set enrichment analysis (GSEA) (Subramanian et al., 2005), leveraging consistency across cell line, dose, and treatment-time contexts so that drugs with reproducible mechanism-level concordance are prioritized.

**Fig. 1.**
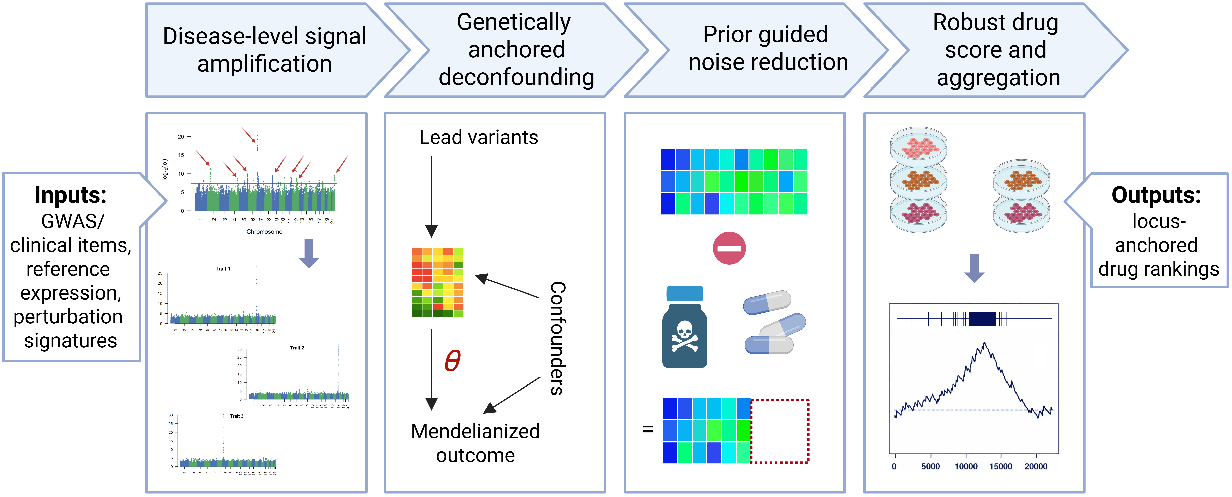
Overview of SIEVE. The algorithm amplifies disease-relevant signal by decomposing complex phenotypes into locus-anchored subphenotypes, genetically calibrating expression-derived mechanisms, removing nonspecific perturbational noise using negative anchors, and aggregating evidence across experimental contexts to produce locus-anchored drug rankings.

Taken together, SIEVE performs signal amplification at several levels: it “Mendelianizes” complex disease phenotypes, deconfounds reference expression, incorporates prior pharmacological knowledge to suppress irrelevant variation, and aggregates evidence across contexts to produce more robust drug-level rankings. Across experiments, SIEVE yields calibrated, genetically grounded perturbagen rankings from high-throughput expression assays and achieves state-of-the-art accuracy.

## Methods

We now elaborate on the SIEVE pipeline in detail.

### Genetically isolating biological mechanisms

We first learn a set of composite outcomes 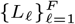 that aim to capture genetically isolated biological mechanisms. To construct these outcomes, we apply the Mendelianization algorithm (Strobl, 2025), which is distinct from Mendelian randomization. Mendelianization takes individual clinical items (e.g., symptoms, laboratory values, and physical examination findings) and combines them into composite outcomes so that, as the number of available items increases, the genetic association for each composite becomes progressively concentrated in fewer loci. In the limit, this concentration collapses to a single locus, and the underlying theory guarantees that at least one variant in the locus is causal for the corresponding composite outcome. In finite settings with only a modest number of items (e.g., ≤ 20), the association may not fully collapse to one locus, but Mendelianization still typically reduces the number of associated loci substantially.

Mendelianization learns one composite outcome per variant. For each candidate variant *i*, it constructs a phenotype *L*_*i*_ by linearly combining the available clinical items to maximize association with the genotype at *i*. Variants for which the corresponding *L*_*i*_ is genome-wide significant define *index variants* and their associated index loci. For each such variant *i*, we then fix *L*_*i*_ and perform a genome-wide association scan using *L*_*i*_ as the phenotype, yielding summary statistics *Ƶ*_*i*_(*j*) over all variants *j*. This scan typically identifies one or a small number of associated loci. For a learned outcome *L*_*ℓ*_, let

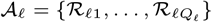

denote the corresponding set of associated loci. We summarize each locus ℛ_*ℓq*_ ∈ 𝒜_*ℓ*_ by its lead variant and retain these lead variants for downstream calibration. In the ideal case *Q*_*ℓ*_ = 1, so *L*_*ℓ*_ is effectively anchored to a single locus; with fewer clinical items, *Q*_*ℓ*_ may be larger.

### Inputs for mechanism scoring: patient and perturbation expression

For a given learned outcome *L*_*ℓ*_, let 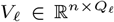 denote the centered and standardized genotype matrix of the retained lead variants across *n* individuals, with one column per locus in 𝒜_*ℓ*_. We also use a drug perturbation compendium such as LINCS L1000 (Subramanian et al., 2017). Rather than working with separate drug and control profiles, we assume a matrix of differential signatures comparing each drug condition with its matched control,

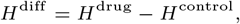

where *H*^diff^ ∈ ℝ^*p×m*^, each column corresponds to one perturbation condition (drug, dose, time, cell line), and each row corresponds to one gene in the perturbation gene universe of size *p*.

### Constructing an outcome-specific, locus-anchored, genetically calibrated mechanism vector

For each learned outcome *L*_*ℓ*_, we construct a calibrated mechanism direction in gene space by combining patient-level genotype and reference-expression data with the Mendelianization summary association statistics for *L*_*ℓ*_. Let *G* ∈ ℝ^*n×p*^ denote the reference-expression matrix for the same individuals aligned across the *p* genes. The downstream calibration step operates at the level of the learned outcome *L*_*ℓ*_, using one lead variant from each associated locus as an instrument.

#### Genetically driven embedding

We first compute the cross-covariance between the lead variants and the gene-wise standardized reference-expression matrix. Let *s* ∈ ℝ^*p*^ denote the vector of gene-wise standard deviations in *G*, and write *G/s* for column-wise scaling of *G* by *s*. We define

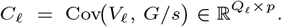

We then compute a singular value decomposition (SVD),

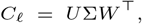

and retain the top *k* = min(*Q*_*ℓ*_, *p*) right singular vectors *W*_*k*_ ∈ℝ^*p×k*^. These vectors define an outcome-specific, locus-anchored low-dimensional mechanism space. We form patient factor scores

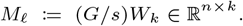

This step extracts expression variation that is maximally coupled, on a gene-wise standardized scale, to the genetic variation represented by the retained lead variants for *L*_*ℓ*_.

#### Calibration to the Mendelianized target

The embedded factors *M*_*ℓ*_ are driven by the genotype matrix *V*_*ℓ*_, which allows us to use the lead variant set *V*_*ℓ*_ as an instrument set for calibrating a mechanism direction in expression space that shifts the Mendelianized outcome *L*_*ℓ*_. In the ideal setting where *L*_*ℓ*_ is observed at the individual level, we would estimate the mechanism direction *θ* ∈ ℝ^*k*^ by two-stage least squares (2SLS) (Basmann, 1957):

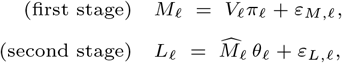

where 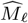 denotes the fitted values from the first stage.

In practice, we do not require individual-level measurements of *L*_*ℓ*_. Instead, we combine (i) individual-level patient data for (*V*_*ℓ*_, *M*_*ℓ*_) with (ii) Mendelianization summary statistics describing association between the lead variants in *V*_*ℓ*_ and *L*_*ℓ*_. After centering and standardizing each column of *M*_*ℓ*_, we estimate the instrument–mechanism association matrix

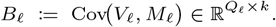

We also form 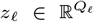, the vector of Mendelianization *z*-statistics for outcome *L*_*ℓ*_ restricted to the lead variants in *V*_*ℓ*_. As the number of outcome items increases, Mendelianization concentrates the genetic signal for each learned outcome into progressively fewer loci, ideally a single locus. In that regime, we will see in the next section that the variant–*L*_*ℓ*_ association pattern is needed only up to scale, because downstream drug scoring depends only on the direction of the resulting gene-weight vector after normalization. Accordingly, we use the local *z*-statistics as the summary-data analogue of the variant–*L*_*ℓ*_ association pattern and estimate *θ*_*ℓ*_ via the ridge-stabilized linear system

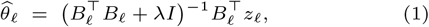

where a small *λ >* 0 improves numerical stability. This estimator therefore recovers the mechanism direction up to an unknown positive scalar factor.

We next map the calibrated direction back to the original expression scale via

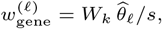

where division by *s* reverses the earlier gene-wise standardization used to construct *C*_*ℓ*_ and *M*_*ℓ*_. This yields a mechanism vector *w*^(*ℓ*)^ ∈ ℝ^*p*^ that represents the calibrated, outcome-specific, locus-anchored gene program through which variation represented by the retained lead-variant set for *L*_*ℓ*_ shifts the Mendelianized target *L*_*ℓ*_.

### Anchor-guided projection in gene space

We often have prior knowledge about drugs that are unlikely to be therapeutically relevant for the disease, such as broadly cytotoxic compounds. We use this information to suppress perturbational directions that are more likely to reflect nonspecific or harmful effects than disease-relevant mechanisms. Specifically, we define a negative anchor set 𝒜_−_ consisting of perturbation conditions corresponding to drugs known to be ineffective or harmful, and we represent this anchor set as a low-rank subspace in gene space by computing a truncated SVD of the perturbation matrix restricted to the corresponding columns:

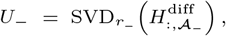

where 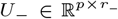 contains the leading left singular vectors (orthonormal columns). By default, we set *r*_−_ = 60, a moderately large value chosen to capture a broad range of nuisance expression directions without spanning most of the perturbational space. We compute this basis once and reuse it for all learned outcomes.

We next refine the calibrated mechanism vector using the negative-anchor subspace. Let Π_*U*_ (*w*) = *U*(*U* ^⊤^*w*) denote orthogonal projection of a vector *w* onto the column space of *U*, and let Π_*U*_ (*H*) = *UU* ^⊤^*H* denote the corresponding column-wise projection of a matrix *H* onto the same gene-space subspace. For each learned outcome *L*_*ℓ*_, we remove the component of the mechanism vector aligned with the negative-anchor subspace:

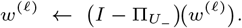

We apply the same refinement to the perturbation signatures:

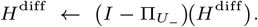

Equivalently, each perturbation signature 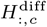 is transformed by the same projection used for *w*^(*ℓ*)^. This projection-based refinement subtracts expression modes associated with negative-control drugs from both the mechanism vector and the perturbation signatures, thereby reducing irrelevant variation before drug scoring.

### Condition-level drug scoring and drug-level aggregation

For each tissue-matched perturbation condition *c*, we score alignment between the perturbation signature and the refined mechanism vector using cosine similarity:

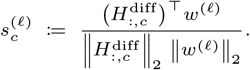

This normalization emphasizes directional concordance rather than signature magnitude. Notice that, since the above score depends on *w*^(*ℓ*)^ only through its direction, the earlier calibration step need only recover *θ*_*ℓ*_ up to a positive scalar factor (Equation (1)).

Because the same perturbagen may appear under multiple experimental contexts (e.g., cell line, dose, and time), we next aggregate condition-level evidence at the drug level using fast gene-set enrichment analysis (FGSEA) (Korotkevich et al., 2016). For a given learned outcome *L*_*ℓ*_, we rank all perturbation conditions by their cosine similarity 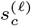. We then treat each drug as a set of its constituent perturbation instances and run FGSEA on this ranked list, yielding a normalized enrichment score (NES) and nominal *p*-value. Intuitively, a large-magnitude NES indicates that a drug’s perturbation instances occur preferentially near one end of the ranked condition list, implying consistent mechanism-level concordance or discordance across contexts. Finally, we rank perturbagens by the nominal p-value.

### Pseudocode and time complexity

We provide pseudocode for SIEVE in Supplementary Materials 5.1. As shown in Supplementary Materials 5.2, the algorithm has quasilinear dependence on the numbers of variants and perturbation conditions and linear dependence on the gene dimensions when viewing the remaining quantities as fixed. By contrast, the runtime includes cubic dependence on the numbers of outcome items and lead variants, but these quantities are small in practice and therefore are unlikely to dominate the overall computational cost.

## Results

### Comparators

We compared SIEVE against the following four algorithms:

1. **DrInsight** (Chan et al., 2019) takes a ranked disease gene-expression signature and compares it to ranked drug-induced expression profiles, identifying genes that show strong opposite-order behavior between disease and drug. It then scores each drug instance with an outlier-sum statistic and uses a K-S test to rank drugs by how strongly their signatures are enriched for reversing the disease pattern.
2. **metaLINCS** (Kwee et al., 2022) computes a gene-level connectivity score between the input disease signature and each individual LINCS perturbation profile, then groups all profiles belonging to the same compound and performs a second-level GSEA-style enrichment test over that grouped set. It outputs a compound-level NES and significance values, so drugs are ranked by how consistently their multiple profiles show disease-reversing connectivity across conditions.
3. **Pathophenotypic Congruity Scores** (PACOS) (Halu et al., 2025) quantify each drug by evaluating how the gene-expression changes it induces relate to those observed in a given disease. Rather than directly comparing the drug and disease signatures, PACOS assesses how each aligns with a large reference panel of disease signatures. A strongly negative PACOS suggests that the drug is likely to reverse the input disease-associated program.
4. **Transcriptome-informed Reversal Distance** (TReD) (He et al., 2025) builds genetically anchored disease signatures, represents both disease signatures and LINCS drug response profiles in a common gene-level vector space, and then defines a reversal distance that quantifies how strongly each drug opposes the disease signature. It uses permutation tests to flag drugs with unusually large, consistent reversal distances across tissues and cell types, thereby prioritizing genetically anchored repurposing candidates with higher chances of true therapeutic benefit.

We also conducted **ablation analyses using three variants of SIEVE**: (1) without Mendelianization, in which we replaced the Mendelianization step with a standard GWAS of a single target phenotype; (2) without the negative anchor set; and (3) without both Mendelianization and the negative anchor set.

### Metrics

We evaluated each method on its ranked output using **precision@***k* and **recall@***k* at *k* = 10, 25, and 50. We also assessed global ranking performance with **normalized mean inverse rank (MIR)**, which measures the average inverse rank of effective drugs after rescaling so that 0 corresponds to random ranking and 1 to the ideal ranking, as well as **average precision** and **area under the precision–recall curve (AUPR)**. In addition, we examined **lift curves** to compare enrichment across the ranked list and recorded **runtime** to assess computational efficiency.

To enable fair comparison across methods that can return different numbers of ranked lists, we used a permutation framework that preserves each method’s output structure. For each method, we first reoriented scores so that larger values always indicated greater evidence of therapeutic relevance in the intended direction. We then organized the reoriented outputs into a drug-by-list score matrix and defined the observed statistic for each drug as the maximum score across all lists in which that drug appeared. To assess significance, we generated an empirical null by permuting score-to-drug assignments within each ranked list, thereby preserving the number of ranked lists, the number of drugs in each list, and each drug’s pattern of list membership. After each permutation, we recomputed the drug-level maximum statistic and estimated an empirical *p*-value as the proportion of permutations in which the null statistic was at least as large as the observed value. We then ranked drugs by increasing empirical *p*-value.

### Simulations

We first conducted simulations by creating realistic drug perturbation, genetic and gene expression data. We created semi-synthetic genetic data by first downloading individual-level autosomal genotype data from European-ancestry participants in the 1000 Genomes Phase 3 dataset (Consortium et al., 2015). After regressing out the first five genetic principal components, we partitioned the genome into 1006 linkage disequilibrium blocks using the optimal LD-splitting algorithm (Privé, 2022) with an *r*^2^ threshold of 0.02 for ignoring pairwise correlations, a window size of 500 kb, a maximum block size of 2000 variants, and a minimum block size of 200 variants. We then sampled 3000 SNPs from these residualized genotypes, selected 3 causal loci from distinct LD blocks, and chose one causal variant per locus; for each locus, we defined a sparse gene program over 1500 genes with support size 80 and unit *ℓ*_2_ norm. Using these locus-anchored programs, we simulated a GWAS cohort of 100,000 individuals and a reference-expression cohort of 500 individuals, with gene expression generated as a linear combination of causal genotype effects, 3-dimensional latent confounding, and independent zero-mean Gaussian noise with standard deviation 0.7.

We then propagated this expression signal into 50 latent phenotypic dimensions and subsequently into 25 observed phenotype items through linear mixing. The same latent confounders that affected expression also affected the phenotypes. Noise in the latent phenotypic dimensions was Gaussian with covariance induced by the latent mapping, whereas noise in the observed phenotype items was independent Gaussian with standard deviation 1. Next, we computed SNP– trait *z*-statistics for each item using per-item GWAS sample sizes drawn uniformly from 50,000 to 100,000, and we defined a total-score phenotype as the first principal component of the observed items. For the perturbational data, we simulated drug signatures across two cell lines with 4 replicates per drug, while varying dose (low, high) and exposure time (6h, 24h); these signatures combined locus-aligned signal, 4 nuisance transcriptional factors, context-specific effects, and independent residual Gaussian noise with standard deviation 0.35. Finally, we instantiated three perturbagen classes: 6 true therapeutics per locus (18 total), 80 negative-anchor compounds, and 120 null compounds, thereby creating a benchmark in which the ground-truth relationships among loci, gene programs, phenotypes, and perturbational responses were known but embedded within realistic confounding and experimental heterogeneity. The simulation code is available in the GitHub repository.

We ultimately simulated 40 paired genetic, gene expression, and drug-perturbation datasets and applied all algorithms to each dataset triplet. Table 1 summarizes the results. SIEVE achieved by far the strongest performance across all evaluation metrics. The lift curves further supported this result, showing that SIEVE consistently remained well above the comparator methods across the ranked list (Figure 2). Ablation analyses likewise showed that both Mendelianization and the negative-anchor component were required for optimal performance (Supplementary Table S1 and Supplementary Figure S1). These findings remained significant after Benjamini– Yekutieli correction (Benjamini and Yekutieli, 2001), with adjusted *p <* 0.05 across all 36 SIEVE-versus-comparator comparisons (4 comparator algorithms *×* 9 metrics) and all 27 ablation comparisons (3 ablations *×* 9 metrics). SIEVE also completed in approximately 2 minutes on average. Taken together, these results indicate that SIEVE was both accurate and computationally practical, and that its major components each contributed materially to its overall performance.

**Table 1.** Performance in the synthetic-data experiments. Entries report the mean *±* standard deviation across simulation replicates.

|  | DrInsight | PACOS | metaLINCS | TReD | SIEVE |
| --- | --- | --- | --- | --- | --- |
| Precision@10 | 0.1000 ± 0.093 | 0.1000 ± 0.084 | 0.1000 ± 0.096 | 0.0725 ± 0.075 | **0.6800** ± 0.168 |
| Precision@25 | 0.0850 ± 0.058 | 0.0990 ± 0.052 | 0.1000 ± 0.055 | 0.0830 ± 0.056 | **0.4530** ± 0.109 |
| Precision@50 | 0.0820 ± 0.036 | 0.0895 ± 0.034 | 0.0940 ± 0.031 | 0.0915 ± 0.037 | **0.2910** ± 0.053 |
| Recall@10 | 0.0555 ± 0.051 | 0.0555 ± 0.047 | 0.0555 ± 0.053 | 0.0402 ± 0.041 | **0.3777** ± 0.093 |
| Recall@25 | 0.1180 ± 0.081 | 0.1375 ± 0.073 | 0.1388 ± 0.076 | 0.1152 ± 0.077 | **0.6291** ± 0.152 |
| Recall@50 | 0.2277 ± 0.101 | 0.2486 ± 0.095 | 0.2611 ± 0.088 | 0.2541 ± 0.103 | **0.8083** ± 0.147 |
| Normalized MIR | 0.0001 ± 0.108 | 0.0120 ± 0.115 | 0.0136 ± 0.107 | -0.0001 ± 0.100 | **0.7187** ± 0.168 |
| Avg. Precision | 0.1042 ± 0.034 | 0.1097 ± 0.033 | 0.1101 ± 0.029 | 0.1054 ± 0.029 | **0.5943** ± 0.146 |
| AUPR | 0.0930 ± 0.031 | 0.0983 ± 0.031 | 0.0991 ± 0.026 | 0.0940 ± 0.025 | **0.5828** ± 0.150 |
| Time (s) | 2.8100 ± 3.555 | 15.501 ± 7.255 | 10.952 ± 7.059 | 7.6970 ± 5.365 | 107.04 ± 34.078 |

**Fig. 2.**
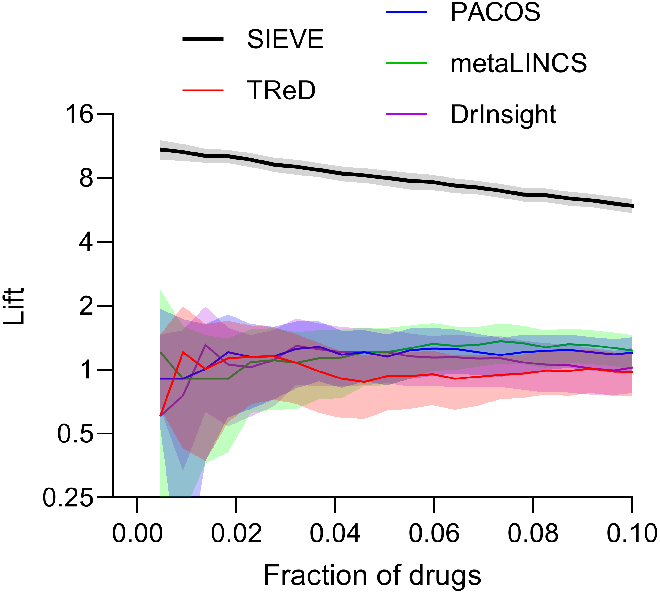
Mean lift curves of algorithms. Shaded bands denote 95% confidence intervals.

### Depression and Generalized Anxiety

We applied SIEVE to summary statistics from the Pan-UK Biobank (Karczewski et al., 2025), focusing on 52 non-collinear Mental Health Questionnaire items capturing lifetime depression and generalized anxiety. As the gene expression reference, we used GTEx v10 data from Brodmann area 9 of the dorsolateral prefrontal cortex (Consortium, 2020). This region has been frequently implicated in major depressive disorder by convergent functional and connectivity studies (Pizzagalli and Roberts, 2022). We then lifted variants from GRCh37 to GRCh38 and harmonized alleles across datasets, retaining only variants shared by both resources with matching reference and alternate allele assignments. We further restricted the analysis to healthy neuronal and neural progenitor cell lines in LINCS L1000. For anchoring, we included a negative set of 218 compounds that were toxic, carcinogenic, or anti-proliferative oncology agents (Supplementary File).

We manually compiled a silver-standard set of 129 compounds with reported clinical or preclinical evidence for treating depression or anxiety and cite the supporting evidence for each compound (Supplementary File). We list all of the compounds that SIEVE mapped to each lead locus in the Supplementary File. Table 2 summarizes the performance of all algorithms. SIEVE achieved the highest accuracy across all metrics except normalized MIR. The lift curve in Figure 3 further supported these findings, showing that SIEVE outperformed the other algorithms across most fractions of ranked drugs. In addition, SIEVE completed in around 4 minutes. Ablation results revealed that both the Mendelianization step and negative anchor set were critical for maintaining strong performance (Supplementary Table S2 and Supplementary Figure S2). Together, these results indicate that SIEVE provides the best accuracy while maintaining a short runtime.

**Table 2.** Performance on the major depression and anxiety dataset. SIEVE achieved the best performance across all accuracy metrics except normalized MIR.

|  | DrInsight | PACOS | metaLINCS | TreD | SIEVE |
| --- | --- | --- | --- | --- | --- |
| Precision@10 | 0.4000 | 0.1000 | 0.4000 | 0.3000 | **0.7000** |
| Precision@25 | 0.2400 | 0.0400 | 0.3200 | 0.3600 | **0.5600** |
| Precision@50 | 0.1600 | 0.0400 | 0.2200 | 0.2400 | **0.4200** |
| Recall@10 | 0.0310 | 0.0114 | 0.0310 | 0.0294 | **0.0542** |
| Recall@25 | 0.0465 | 0.0114 | 0.0620 | 0.0882 | **0.1085** |
| Recall@50 | 0.0620 | 0.0229 | 0.0852 | 0.1176 | **0.1627** |
| Normalized MIR | 0.0678 | 0.1599 | **0.3738** | 0.1815 | 0.3337 |
| Avg. Precision | 0.0922 | 0.0956 | 0.1373 | 0.1387 | **0.2117** |
| AUPR | 0.0953 | 0.0973 | 0.1399 | 0.1450 | **0.2167** |
| Time (s) | 595.73 | 4093.2 | 224.08 | 161.35 | 234.40 |

**Fig. 3.**
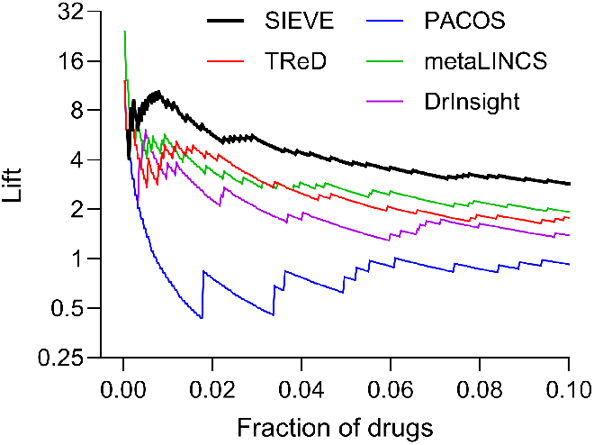
Lift curve for the major depression and anxiety dataset. SIEVE achieved higher lift than the comparator methods across most top screening fractions.

### Alcohol Use Disorder

We next applied the algorithms to a more challenging setting: alcohol use disorder (AUD), for which few therapeutic compounds have been identified, even at the preclinical stage. We downloaded summary statistics for the 10 individual items of the Alcohol Use Disorders Identification Test (AUDIT) from the Pan-UK Biobank (Karczewski et al., 2025). Because the outcome space is limited, we did not expect Mendelianization to localize signal as effectively as in prior analyses. Even so, we hypothesized that SIEVE would maintain strong performance despite the reduced ability to localize each learned outcome to an approximately single causal locus. We paired these data with GTEx v10 expression profiles from the nucleus accumbens (Consortium, 2020), a brain region repeatedly implicated in alcohol use (Bracht et al., 2021), as well as with neural and neural progenitor cell lines. We also manually curated a set of 37 candidate compounds with reported clinical or preclinical evidence for treating AUD and cite the supporting evidence for each compound (Supplementary File). Finally, we used the same negative anchor set of 218 compounds as in the previous application.

We report all compounds that SIEVE mapped to each lead locus in the Supplementary File, and Table 3 summarizes the corresponding accuracy results. SIEVE delivered the strongest performance across all metrics. The lift plots further supported this advantage: SIEVE maintained the highest curve across most fractions of ranked drugs (Figure 4). In the ablation analysis, the negative anchor set proved critical for achieving maximal performance, whereas Mendelianization did not consistently improve results, as expected given the availability of only 10 AUDIT items (Supplementary Table S3 and Supplementary Figure S3). SIEVE also completed the analysis in under 4 minutes. Overall, the AUD results closely mirrored those for depression and anxiety: despite the limited number of outcomes, SIEVE achieved the strongest performance while preserving practical computational efficiency.

**Table 3.**
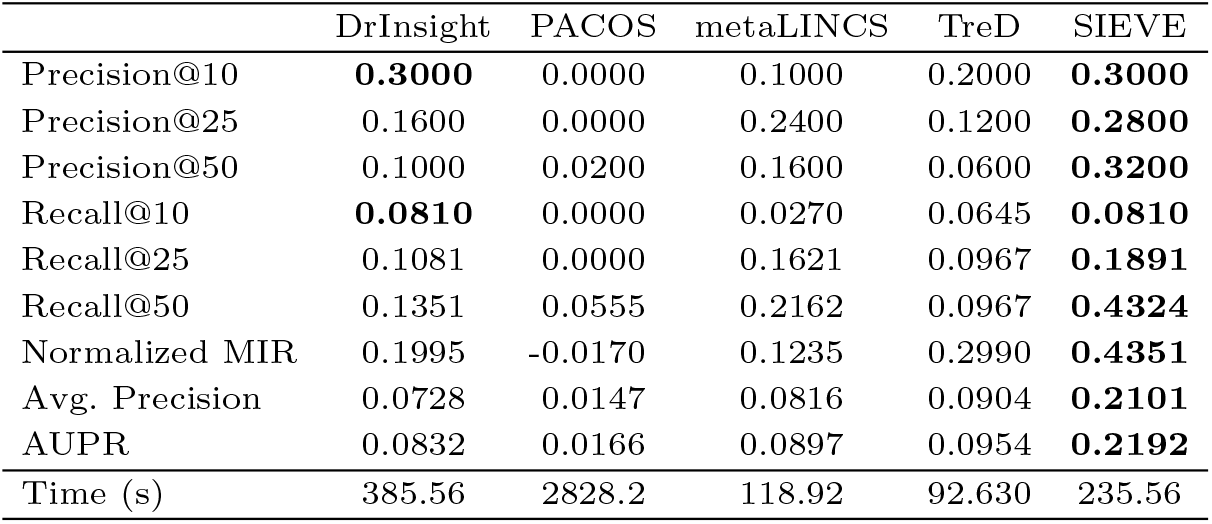
Performance on the alcohol use disorder dataset. SIEVE again achieved the best performance across all accuracy metrics.

|  | DrInsight | PACOS | metaLINCS | TReD | SIEVE |
| --- | --- | --- | --- | --- | --- |
| Precision@10 | **0.3000** | 0.0000 | 0.1000 | 0.2000 | **0.3000** |
| Precision@25 | 0.1600 | 0.0000 | 0.2400 | 0.1200 | **0.2800** |
| Precision@50 | 0.1000 | 0.0200 | 0.1600 | 0.0600 | **0.3200** |
| Recall@10 | **0.0810** | 0.0000 | 0.0270 | 0.0645 | **0.0810** |
| Recall@25 | 0.1081 | 0.0000 | 0.1621 | 0.0967 | **0.1891** |
| Recall@50 | 0.1351 | 0.0555 | 0.2162 | 0.0967 | **0.4324** |
| Normalized MIR | 0.1995 | -0.0170 | 0.1235 | 0.2990 | **0.4351** |
| Avg. Precision | 0.0728 | 0.0147 | 0.0816 | 0.0904 | **0.2101** |
| AUPR | 0.0832 | 0.0166 | 0.0897 | 0.0954 | **0.2192** |
| Time (s) | 385.56 | 2828.2 | 118.92 | 92.630 | 235.56 |

**Fig. 4.**
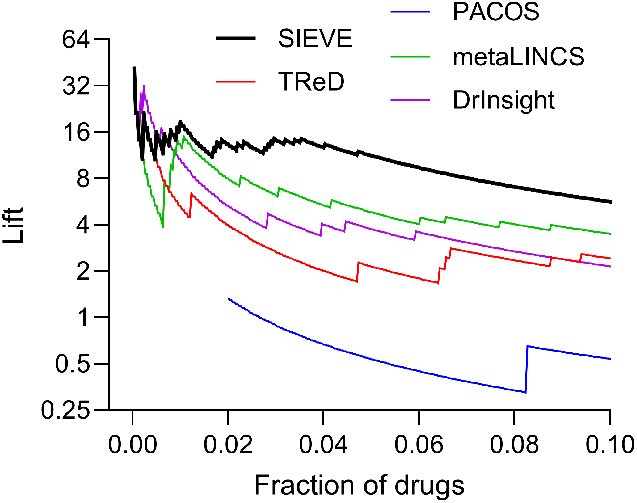
Lift curve for the alcohol use disorder dataset. SIEVE again achieved higher lift than the comparator methods across most screening fractions up to 0.10.

## Discussion

We introduced SIEVE, a method that addresses both phenotypic and molecular heterogeneity by decomposing complex disorders into genetically coherent components and amplifying mechanism-relevant signal in reference and perturbational expression data through the joint use of genetics, instrumental variable analysis, and prior knowledge. Across both synthetic and real datasets, SIEVE outperformed existing comparators. Ablation analyses further showed that the framework’s most novel components contributed meaningfully to overall performance. Taken together, these results suggest that resolving heterogeneity at both the phenotype and molecular levels is critical for accurate drug prioritization in complex disorders.

The magnitude and consistency of SIEVE’s improvement likely reflect the intrinsic difficulty of the task: therapeutic signal is masked by phenotypic heterogeneity, experimental variation, and molecular noise. Recovering that signal appears to require a coordinated, multi-stage strategy rather than methods that address only isolated aspects of the problem. Our results therefore suggest that comprehensive modeling of these distinct but interacting sources of variation is necessary to achieve strong performance.

An additional strength of SIEVE is that it achieves signal amplification through data-adaptive statistical modeling rather than relying solely on conventional preprocessing. Related ideas have also been used in clinical data analysis (Strobl, 2026), but the underlying principle is general and likely helps the method accommodate dataset-specific sources of noise and latent confounding. More broadly, these results suggest that data-adaptive approaches to noise mitigation and confounding control can substantially improve performance in high-dimensional drug-prioritization settings.

SIEVE nevertheless has several limitations. First, the disease-decomposition step is most effective when multiple outcomes are available, as this allows Mendelianization to construct composite traits that more closely approximate Mendelian mechanisms. In some complex disorders, however, the available phenotypic information is limited to coarse diagnostic labels or aggregate severity measures, which reduces this advantage. In such settings, one practical alternative is to begin with loci identified by standard GWAS and choose *k* to be substantially smaller than the number of associated loci, thereby enforcing a lower-dimensional mechanistic representation. Second, SIEVE prioritizes drugs at the locus level rather than at the level of the individual patient. We were primarily interested in addressing the foundational problem of improving performance in the population setting, but future work should extend the framework to recommend patient-specific therapies based on each individual’s genetic profile. Third, the method depends on perturbational data from biologically relevant cell lines. In practice, available profiles may come only from unrelated or suboptimal cellular contexts, which may attenuate performance. Methods that infer or approximate disease-relevant perturbational responses from imperfectly matched cell lines would therefore increase the practical utility of the framework (e.g., (Sapashnik et al., 2023; Iwata et al., 2019)). Overall, future work should focus on extending SIEVE to settings with limited phenotypic resolution, enabling patient-specific therapeutic prioritization, and improving robustness to non-ideal cellular models.

## Data Availability

All data are openly available and include LINCS L1000 perturbation data, GTEx variant data, GTEx expression data, Pan-UK Biobank summary statistics, and 1000 Genomes Phase 3 data.

https://clue.io/data/CMap2020#LINCS2020

https://gtexportal.org/home/protectedDataAccess

https://gtexportal.org/home/downloads/adult-gtex/bulk_tissue_expression

https://pan.ukbb.broadinstitute.org/downloads

https://ftp.1000genomes.ebi.ac.uk/vol1/ftp/release/20130502/

## Supplementary Materials

### Pseudocode

We present the pseudocode for SIEVE in Algorithm 1. The algorithm first applies Mendelianization to the matrix of outcome-item *z*-statistics in Line 1 in order to identify learned outcomes and obtain the corresponding Mendelianization scans *Ƶ*. It then restricts the perturbational data to the retained cell lines and constructs a low-rank basis *U*_−_ from the negative-anchor perturbations in Line 3. This basis defines the nuisance subspace that SIEVE removes from both the mechanism vector and the perturbational signatures, while the corresponding projected condition norms *ν*_*c*_ in Line 4 provide the denominators for the subsequent condition-level cosine-similarity scores.

Next, for each learned outcome *L*_*ℓ*_, SIEVE performs a genetically driven embedding by computing the covariance between the local lead variants *V*_*ℓ*_ and the scaled reference expression matrix *G/s* in Line 8, followed by a truncated singular value decomposition in Line 9. The resulting right singular vectors define an outcome-specific, locus-anchored low-dimensional mechanism space. SIEVE then calibrates this embedded representation to the Mendelianized target by regressing the Mendelianization *z*-statistics for *L*_*ℓ*_ restricted to the lead variants in *V*_*ℓ*_ onto the instrument–mechanism association matrix *B*_*ℓ*_ through a ridge-regularized calibration step, which yields the coefficient vector *θ*_*ℓ*_ in Line 15. Mapping *W*_*ℓ,k*_*θ*_*ℓ*_*/s* back to gene space produces an outcome-specific, locus-anchored mechanism vector *w*_*ℓ*_ in Line 16, which is subsequently projected away from the negative-anchor subspace in Line 18 and normalized. Using this refined mechanism vector, the algorithm computes a condition-level cosine-similarity score 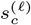 for every retained perturbation condition *c* in Line 21.

SIEVE finally aggregates these condition-level scores to the drug level by ranking perturbation instances according to 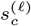, grouping instances by drug identity, and applying FGSEA (Korotkevich et al., 2016) in Line 26 to test whether a drug’s perturbation instances are enriched near one end of the ranked list. This yields learned-outcome-specific normalized enrichment scores, permutation-based nominal *p*-values, and final drug rankings as desired.

#### Algorithm 1 SIEVE

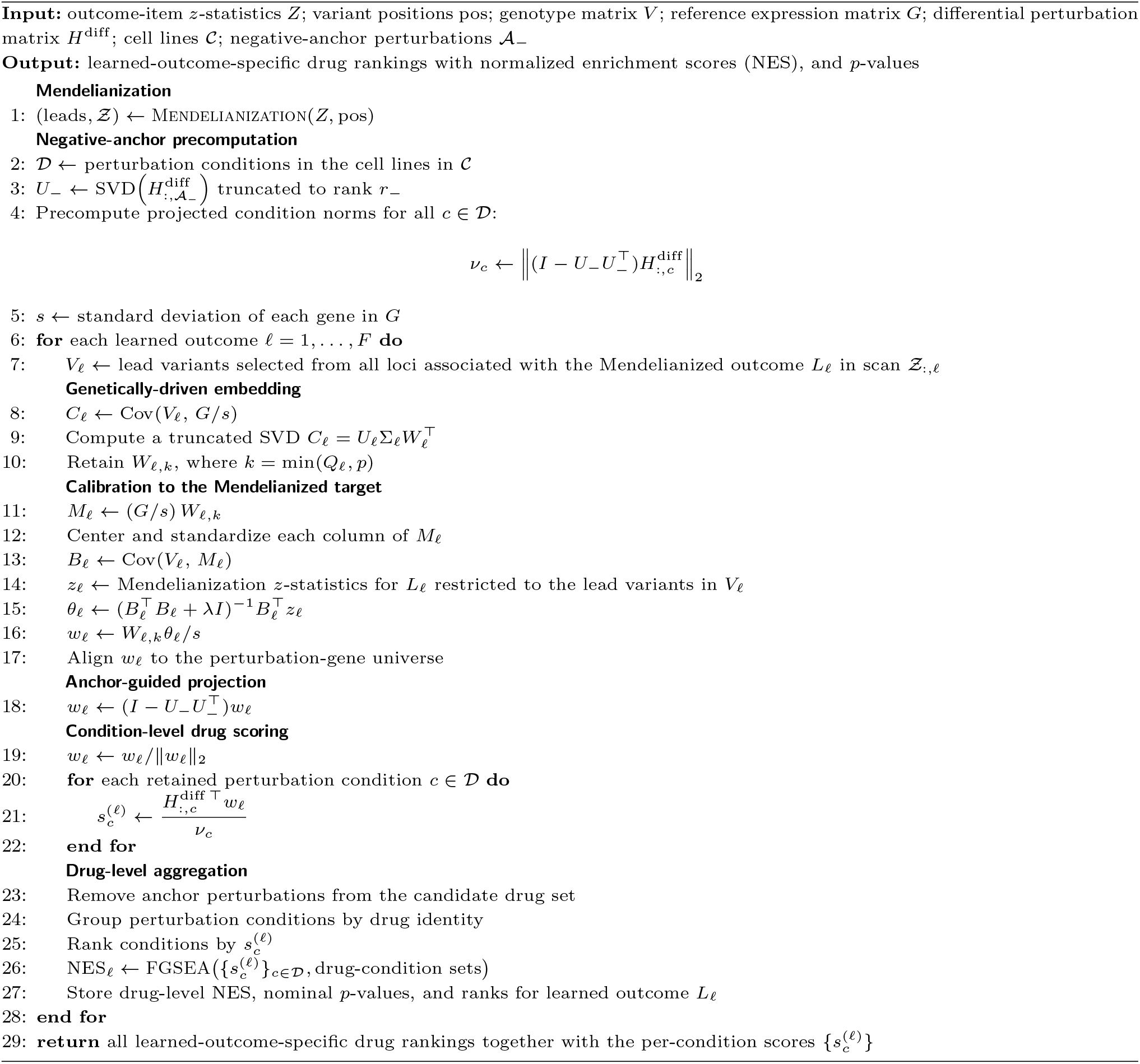

### Computational Complexity Analysis

We now analyze the computational complexity of SIEVE. For simplicity, the Methods described the reference-expression and perturbation gene dimensions using the same symbol *p*; here, we allow them to differ, writing *g* for the number of reference-expression genes and *p* for the number of perturbation genes. Let *h* denote the number of variants entering Mendelianization, *t* the number of clinical items, and *n* the number of reference individuals, *m* the number of perturbation conditions retained after cell-line filtering, and *F* the number of learned outcomes retained after Mendelianization. For learned outcome *L*_*ℓ*_, let *Q*_*ℓ*_ denote the number of lead variants retained across the loci associated with *L*_*ℓ*_ and used to construct the outcome-specific, locus-anchored mechanism. We further let *a*_−_ and *r*_−_ denote the number of negative-anchor conditions and the retained rank of the corresponding negative-anchor basis, respectively.

The total runtime decomposes into four stages: Mendelianization, negative-anchor precomputation, outcome-specific, locus-anchored mechanism construction and scoring, and drug-level aggregation by enrichment analysis. The Mendelianization stage requires sorting the variant positions, which costs *O*(*h* log *h*); forming the phenotype covariance matrix, which costs *O*(*ht*^2^) ; inverting the corresponding *t × t* system, which costs *O*(*t*^3^) ; computing the canonical coefficients, which also costs *O*(*ht*^2^) ; and generating lead-specific *z*-statistics, which costs *O*(*F*(*ht* + *t*^2^)). Thus, after combining terms, this stage costs

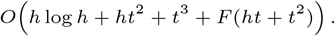

The anchor stage requires constructing the negative-anchor basis by truncated singular value decomposition, which costs *O*(*pa*_−_*r*_−_), and precomputing the corresponding perturbation-space coefficients for the retained conditions, which costs *O*(*pmr*_−_). Thus, after combining terms, this stage costs

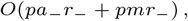

where *r*_−_ is typically small.

For each learned outcome *L*_*ℓ*_, SIEVE requires computing the genotype–expression cross-covariance matrix, which costs *O*(*ngQ*_*ℓ*_); performing a singular value decomposition of the resulting *Q*_*ℓ*_ *× g* matrix, which costs 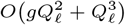 ; forming the embedded factor scores and the instrument–mechanism covariance matrix, which together cost 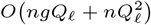 ; solving the ridge-stabilized normal equations, which costs 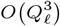 ; projecting the resulting mechanism vector away from the negative-anchor subspace, which costs *O*(*pr*_−_); and computing the condition-level scores across all retained perturbation conditions, which costs *O*(*pm*). Thus, after combining terms and absorbing constant factors, the corresponding per-outcome cost is

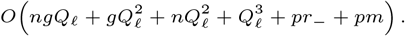

After computing the condition-level scores, SIEVE ranks the *m* retained perturbation conditions, which costs *O*(*m* log *m*), and then performs one FGSEA analysis per learned outcome to aggregate condition-level evidence to the drug level. Let *D*_*ℓ*_ denote the number of retained drugs for learned outcome *L*_*ℓ*_, let *K*_*ℓ*_ denote the maximum number of retained perturbation conditions associated with any one retained drug for learned outcome *L*_*ℓ*_, and let *n*_perm_ denote the number of FGSEA permutations. Using the complexity of FGSEA-simple reported in (Korotkevich et al., 2016), the enrichment step contributes 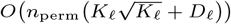 per learned outcome. Therefore, the total drug-level aggregation cost per learned outcome is

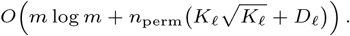

Accordingly, the total runtime can be written as

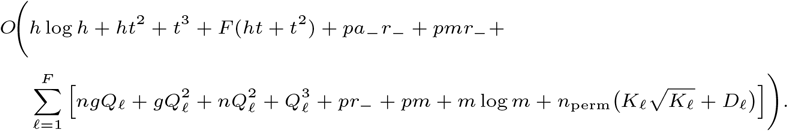

SIEVE therefore scales as *O*(*h* log *h*) in the number of variants and *O*(*m* log *m*) in the number of retained perturbation conditions, while remaining linear in the gene dimensions *g* and *p*. By contrast, the dependence on the number of outcome items *t* and the number of lead variants *Q*_*ℓ*_ includes cubic terms, but these quantities are typically small in practice and therefore do not dominate the runtime.

### Extra Synthetic Data Results

**Table S1.**
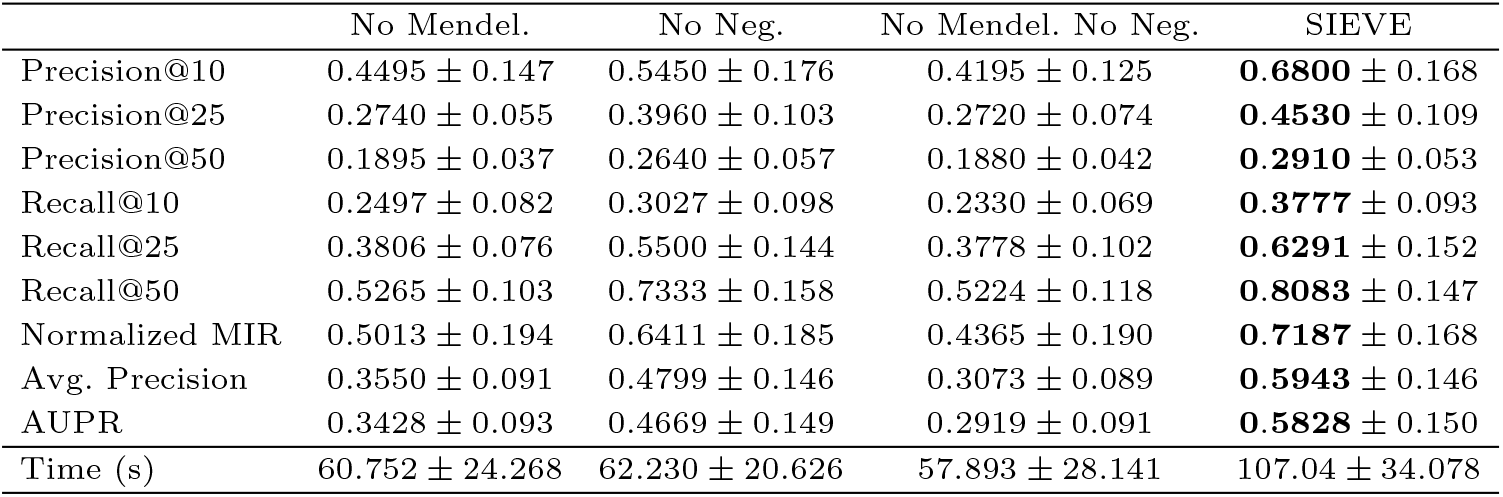
Ablation results for the synthetic datasets. Both the Mendelianization step and negative anchor set were important for maintaining strong performance.

**Fig. S1.**
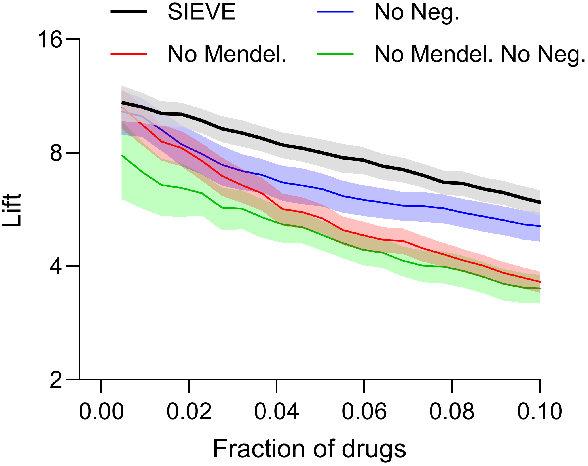
Lift curves for the ablation experiments. SIEVE achieved the strongest performance across most drug fractions.

### Extra Depression and Generalized Anxiety Results

**Table S2.** Ablation results for the depression and anxiety dataset. Both the Mendelianization and negative anchor set steps were critical for maintaining strong performance on this dataset.

|  | No Mendel. | No Neg. | No Mendel. No Neg. | SIEVE |
| --- | --- | --- | --- | --- |
| Precision@10 | 0.2000 | 0.4000 | 0.2000 | **0.7000** |
| Precision@25 | 0.2000 | 0.4000 | 0.1200 | **0.5600** |
| Precision@50 | 0.1800 | 0.3800 | 0.0800 | **0.4200** |
| Recall@10 | 0.0155 | 0.0310 | 0.0155 | **0.0542** |
| Recall@25 | 0.0387 | 0.0775 | 0.0232 | **0.1085** |
| Recall@50 | 0.0697 | 0.1472 | 0.0310 | **0.1627** |
| Normalized MIR | 0.0788 | **0.4238** | 0.1983 | 0.3337 |
| Avg. Precision | 0.1140 | 0.1702 | 0.1083 | **0.2117** |
| AUPR | 0.1176 | 0.1738 | 0.1104 | **0.2167** |
| Time (s) | 192.45 | 276.73 | 321.23 | 234.40 |

**Fig. S2.**
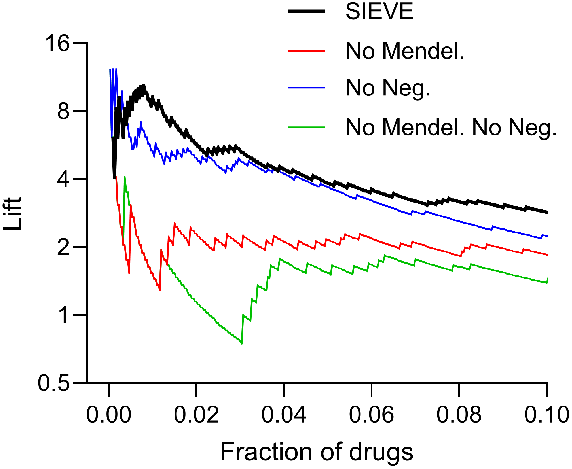
Lift curves for the depression and anxiety dataset.

### Extra Alcohol Use Disorder Results

**Table S3.** Ablation results for the alcohol use disorder dataset experiments. The negative anchor set was critical for maintaining performance on this dataset.

|  | No Mendel. | No Neg. | No Mendel. No Neg. | SIEVE |
| --- | --- | --- | --- | --- |
| Precision@10 | **0.6000** | 0.0000 | 0.1000 | 0.3000 |
| Precision@25 | **0.4400** | 0.0000 | 0.0800 | 0.2800 |
| Precision@50 | 0.2600 | 0.0200 | 0.0400 | **0.3200** |
| Recall@10 | **0.1621** | 0.0000 | 0.0270 | 0.0810 |
| Recall@25 | **0.2972** | 0.0000 | 0.0540 | 0.1891 |
| Recall@50 | 0.3513 | 0.0270 | 0.0540 | **0.4324** |
| Normalized MIR | 0.4034 | -0.0105 | 0.0293 | **0.4351** |
| Avg. Precision | **0.2154** | 0.0318 | 0.0431 | 0.2101 |
| AUPR | **0.2317** | 0.0337 | 0.0471 | 0.2192 |
| Time (s) | 364.34 | 256.26 | 220.39 | 235.56 |

**Fig. S3.**
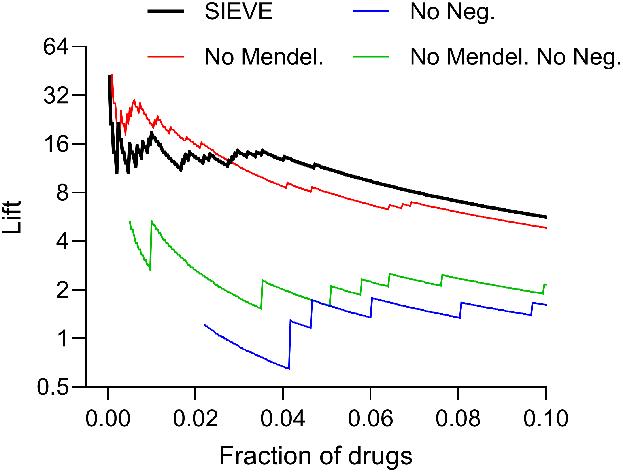
Lift curves for the alcohol use disorder dataset.

